# Prevalence and factors associated with utilization of family planning methods among youth in northwestern Tanzania

**DOI:** 10.64898/2026.02.27.26347237

**Authors:** Upendo Shorusaeli Safari, Leah Anku Sanga, Catherine Massey Safari, Rune Nathaniel Philemon, Jane Januaris Rogathi, Geofrey Nimrod Sigalla

## Abstract

**Introduction:** Youth are challenged globally by sexual and reproductive health problems such as unwanted pregnancy, unsafe abortion and sexually transmitted infections. While family planning methods are a safe and effective way for individuals to responsibly control their sexual and reproductive needs, their use amongst sexually active youth is very low. Of urgency, is to understand factors influencing youth to adopt family planning methods so as to inform future strategies aiming at increasing the use of modern methods of contraception. This study aimed at determining the prevalence and factor associated with utilization of family planning methods among youth in northwestern Tanzania.

**Materials and Methods:** A cross sectional analytical study was conducted among youth aged 15-24 years in selected secondary schools and university Colleges in Nyamagana district, Mwanza Tanzania. Participating institutions and the participants were selected using a multistage sampling method. Data was collected using self-administered questionnaires and later analyzed using SPSS version 25. Univariate and multivariable logistic regression was used to analyze factors associated with the use of family planning; with significance considered at p-value<0.05.

**Results:** A total of 349 participants were enrolled. The prevalence of utilization of family planning methods among sexually active youth was 83.2%. Factors associated with FP use were being female (AOR 2.84; CI: 1.05, 7.67) and not having a peer who is using the method (AOR 0.31; 95% CI: 0.12, 0.82). Poor awareness on availability of FP services at nearby facility was found to be significant (cOR 0.38; 95% CI: 0.16-0.90) during crude analysis but become insignificant when adjusted for other factors

**Conclusion:** Majority of sexually active youth were utilizing FP methods. Sex and peer pressure were significantly associated with family planning use. Therefore, initiatives for advocating comprehensive sexuality education and strengthening youth friendly health clinics are highly proposed to increase consistent contraceptive use among youth.

## Introduction

Youth contribute to a large extent in making the global population. It is documented that 1.8 billion of the total population are youth, whereby 90% live in Low- and middle-income countries (LMIC) (1). According to Tanzanian census in 2022, a total of 11.8 million people were youth, this is equivalent to 19.2% of the entire population of 61.7 million (2). This group of population is challenged by increasing sexual and reproductive health problems such as unwanted pregnancy, unsafe abortion, sexual transmitted infections (3).

Unwanted pregnancies are major public health problem which ends in life threatening conditions in most cases. Worldwide, almost 12 million young women aged 15-19 years give birth each year equivalent to 9% of the total 134 million births in 2024, majority of which occur in Sub-Sahara Africa (4,5). Globally, pregnancy related complications are the leading cause of death among young women aged 15-19 years; their babies are more likely to die as compared to older women (5). The majority of youths are at increased risk of maternal mortality due to pregnancy related complications (6). In Tanzania, about 22% of women aged 15 -19 years have ever been pregnant, and majority of which were unplanned (7). Unsafe abortion is a significant cause of maternal deaths in Tanzania, which in turn contributes to higher maternal mortality rate worldwide (4).

Family planning is one of the safe and effective ways for individuals to responsibly control their sexual and reproductive needs (3). It is documented that the higher fertility rate among young women is a result of low use of modern contraceptives. Despite of low use, family planning methods remain to be of paramount importance in averting unwanted pregnancy and unsafe abortion, and also prevent maternal and children death.

Tanzania Government’s Development Vision 2050 commitment in attaining a prosperous, just, inclusive and self-reliant nation is clear and directional, with goal in health being ensuring people live high quality life and well-being (8). Tanzania Zanzibar’s Development vision 2050 nests on sustainable and inclusive human development supported by reliable and sustainable social services for all (9). Ministries of Health (both mainland and in Zanzibar) have instituted a specific directorate on Reproductive, Maternal and Child health to oversee the advancement of equitable access to sexual reproductive health inclusive of family planning. Scale up of family planning services has always been a top priority by Tanzania government, and efforts are guided by several policies and implementation frameworks such as the FP2030 country commitment, the National Family Planning Costed Implementation Plan, the Health Sector Strategic Plan, and the National Family Planning Procedure Manual, which together provide strategic, operational, and technical direction for equitable access to services (10–13). With contribution from various partners, family planning services are generally considered provided for free with exception to some private facilities where user fee is charged for procedures but not FP commodities. The services are provided through multi-pronged approaches inclusive of facility based and outreach, usually coupled with health information, education and demand generation. Despite these recommendable efforts, prevalence of utilization of modern family planning methods in Tanzania have remained low, especially youths.

The country committed through FP2030 initiative of increasing access and utilization of modern contraceptives among adolescent to 20% and the modern contraceptive prevalence rate (mCPR) of 42% by 2025 have never been met (10). The results of TDHS 2022 showed that mCPR was 31%, a significant stride from the prevalence of 32% registered in 2015; and the utilization of modern family planning among adolescents was 15% compared to 13% (7,14). To contribute to these efforts, it is important to retrieve evidences useful in understanding the prevalence of family planning use and the factors influencing its use among youth. More information from research including the present to provide data that will act as a benchmark on measurement of evaluation for national youth policy implementation and contributing towards improving strategies for implementation of youth friendly reproductive health clinics. By so doing this will tackle their reproductive needs and enhance meeting their reproductive goals.

## Materials and Methods

### Study setting and design

This cross-sectional analytical study was carried out in Nyamagana district, in north-western Tanzania. The district has a population of 363,452, and the major occupations of the residents are agriculture and fishing. The study population included youth aged 15-24 years in Pamba and Mwanza secondary schools and Tandabui College located in the district. All youth who consented to participate in the study were included with exception to those who had stayed for less than six months in the district.

### Sampling size determination and sampling procedure

The minimum sample size in this study was calculated by using prevalence of contraceptive use among youth in Tanzania, where a formula for single population proportion was applied, 95% level of significance and 5% margin error. The sample size used was 349.

To increase the level of efficiency of sampling and composition of individuals in different age groups to be included, a multistage sampling technique was employed.

Youth to participate in this study were hosted within schools and universities, which were treated as clusters and therefore a cluster sampling technique was used in the first stage. A total of two schools and one college were selected through simple random sampling technique using balloting method. Lastly, in order to recruit study participants, systematic random sampling technique was applied whereby sampling interval was calculated proportionately basing on secondary school and college population to determine the sample size per each.

Sampling interval was calculated per school/college by dividing the total number of students in level (N) by sample size (n). Then first participant was selected using lottery method thereafter the remaining participants were selected following regular interval calculated until the required sample size obtained.

### Data collection tools and technique

A self-administered standardized questionnaire with closed-ended questions was used to collect information required from participants in February and March 2018. This was adopted from a study done by Solomon (2014) in Addis Ababa Ethiopia and modified to suit the objectives of this study. The questionnaire consisted of four parts; social demographic characteristics, socio-cultural, awareness of family planning methods and health service related questions.

Prior to the study, pilot study was conducted; the questionnaire was pretested to 30 participants from Kwimba district (youth in similar settings). A pretested self-administered questionnaire in Swahili language, translated from English was distributed among 349 participants who consented to participate in study.

### Data analysis

Data was analyzed using SPSS version 25 after data cleaning. Descriptive statistics were done to summarize mean and standard deviation for continuous variable and frequency and proportion for categorical variables, Univariate and multivariable logistic regression was used to analyze factors associated with the use of family planning. At univariate analyses, significance was considered at p-value≤0.2. Variables which were significant at univariate model were taken to multivariable model. At multivariable model, significance was considered at p-value<0.05. Odds ratios with respective 95% confidence intervals were presented for both univariate and multivariable model.

### Ethical consideration

Ethical clearance for conducting the study was obtained from Kilimanjaro Christian Medical University College Research Ethics and Review Committee (CRERC) with certificate number 2233. Permission to conduct the study was requested and granted from Nyamagana District Executive Officer, and director from TANDABUI institute of health and allied sciences with information sent to participating schools. Written informed consent was obtained from every study participant. Confidentiality was highly observed by ensuring that the participants use code number instead of their names. The participants were well informed that they have freedom to refuse from participation to the study or refrain from answering certain question that doesn’t want to, without any penalty.

## RESULTS

### Social demographic characteristics of study participants

A total of 349 secondary and university college students were enrolled in this study. Participants enrolled in the study were aged from 15 to 24 years with mean age (Standard deviation) of 19.7(1.9) years (Table 1). Majority of participants (55.0%) were aged between 19-21 years. Of the students, 193 (55.3%) were females, one hundred and ninety-five (55.9 %) were participants from secondary schools and more than half of all participants (52.7%) were Roman Catholic followed by 107 (30.7%) who were Christians from other denominations.

**Table 1:** Socio-demographic characteristics of participants (n=349)

| **Variable** |  | **Number (%) of participants** |
| --- | --- | --- |
| **Age (year)** |  |  |
|  | Mean age (SD) | 19.7 (1.9) |
|  | Range (min-max) | 15-24 |
|  | 15-18 | 85 (24.4) |
|  | 19-21 | 192 (55) |
|  | 22-24 | 72 (20.6) |
| **Sex** |  |  |
|  | Male | 156 (44.7) |
|  | Female | 193 (55.3) |
| **Education level** |  |  |
|  | Secondary | 195 (55.9) |
|  | University college | 154 (44.1) |
| **Religion** |  |  |
|  | Roman Catholic | 184 (52.7) |
|  | Other Christians* | 107 (30.7) |
|  | Muslims | 58 (16.6) |

\*Other Christians include denominations such as FPTC, EAGT, AICT, Lutheran, Menonnite and Anglican.

### Utilization of modern family planning methods

Out of 349 participants enrolled in the study, 184 (53%) reported to have ever had sex, majority being females, aged between 19 - 21 years and studying at university level. Among participants who reported to have ever had sex, 153 (83.2%) reported to have used family planning methods. Condom was the commonest family planning method ever used by more than three quarter of participants who have ever had sex (76%) (Figure 1). Other participants reported to have used more than one method such as pills, injectables and implants. Few participants used oral pills, injectables and IUCD (8%, 3%, 2%) respectively.

**Figure 1:**
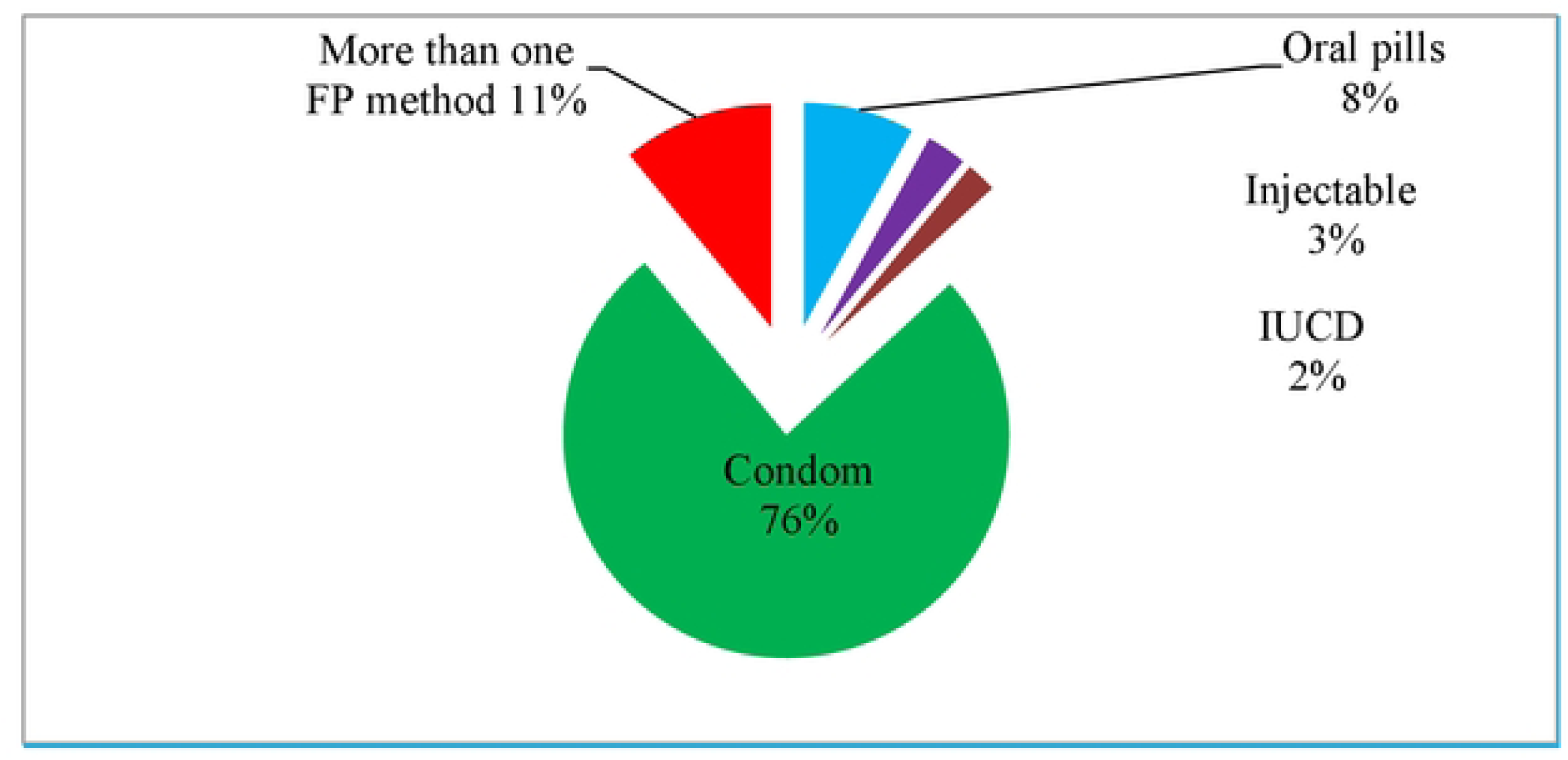
Commonly used family planning methods (n=l53)

Table 2 below describes the characteristics of participants who used family planning methods. The majority of those who used family planning methods 86 (56.2 %) were aged 19 – 21 years, were males (51.0 %) and were from university college (60.1%). Also, the majority of participants (83.4 %), reported that their culture doesn’t prohibit family planning methods use and that their families supported family planning use (50.3%). Majority (83.7%) reported that money was not a hindrance towards family planning use and 93 (61.2%) had friends who had used family planning methods before. On the other hand, almost all of participants who had ever had sex (97.4 %) heard about family planning and 140 (92.1%) knew where to get family planning services including 122 (80.3%) who pointed out that family planning services were available at nearby facilities.

**Table 2:**
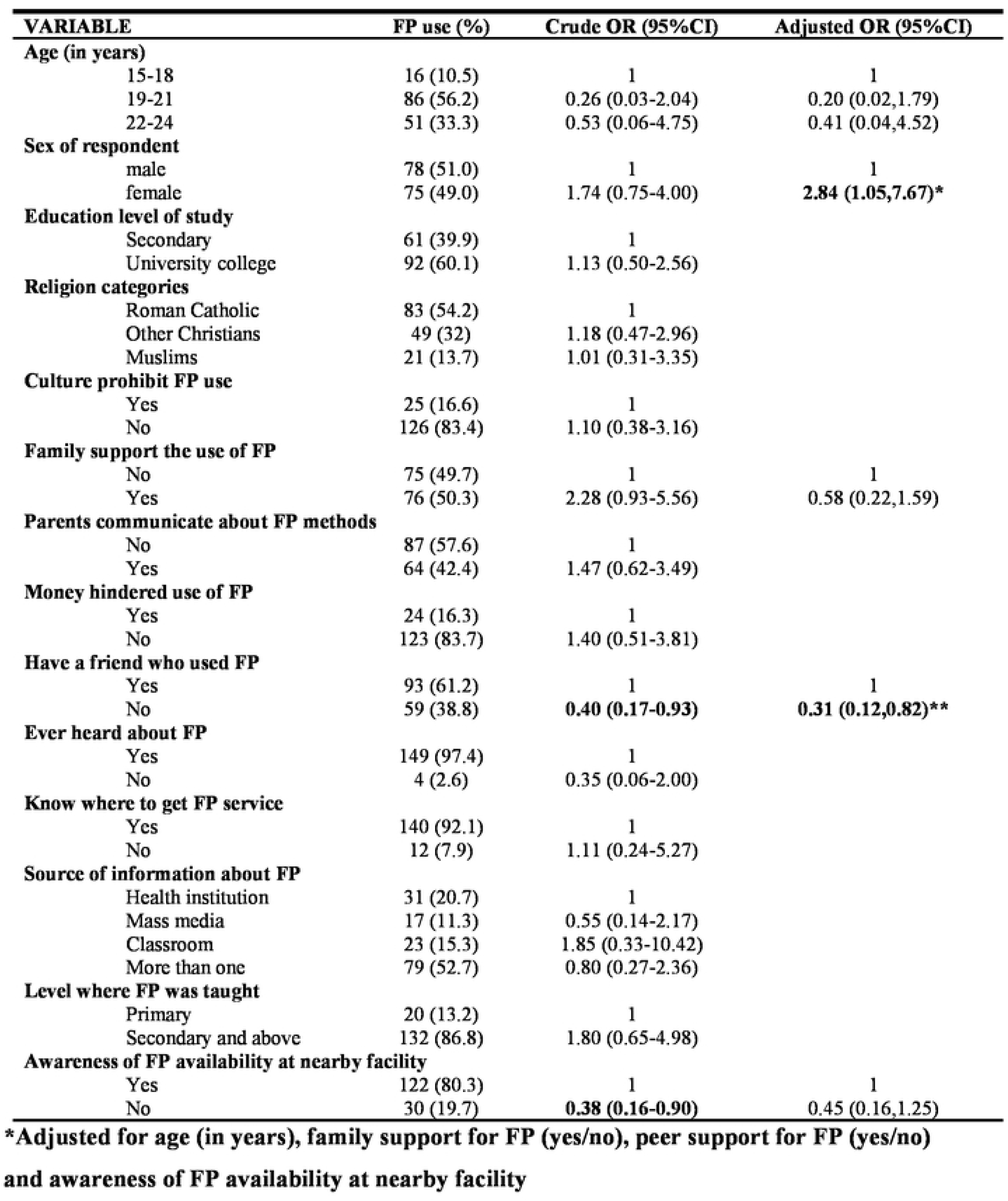
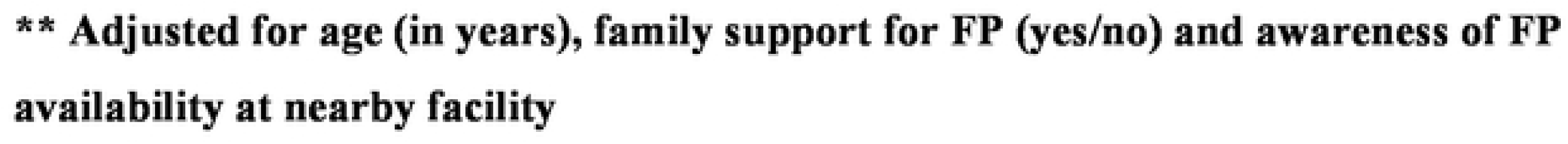
Factors associated with use of family planning among youth in north western Tanzania (n=l84)

| **VARIABLE** | **FP use (%)** | **Crude OR (95%CI)** | **Adjusted OR (95%CI)** |
| --- | --- | --- | --- |
| **Age (in years)** |  |  |  |
| 15-18 | 16 (10.5) | 1 | 1 |
| 19-21 | 86 (56.2) | 0.26 (0.03-2.04) | 0.20 (0.02,1.79) |
| 22-24 | 51 (33.3) | 0.53 (0.06-4.75) | 0.41 (0.04,4.52) |
| **Sex of respondent** |  |  |  |
| male | 78 (51.0) | 1 | 1 |
| female | 75 (49.0) | 1.74 (0.75-4.00) | **2.84 (1.05,7.67)\*** |
| **Education level of study** |  |  |  |
| Secondary | 61 (39.9) | 1 |  |
| University college | 92 (60.1) | 1.13 (0.50-2.56) |  |
| **Religion categories** |  |  |  |
| Roman Catholic | 83 (54.2) | 1 |  |
| Other Christians | 49 (32) | 1.18 (0.47-2.96) |  |
| Muslims | 21 (13.7) | 1.01 (0.31-3.35) |  |
| **Culture prohibit FP use** |  |  |  |
| Yes | 25 (16.6) | 1 |  |
| No | 126 (83.4) | 1.10 (0.38-3.16) |  |
| **Family support the use of FP** |  |  |  |
| No | 75 (49.7) | 1 | 1 |
| Yes | 76 (50.3) | 2.28 (0.93-5.56) | 0.58 (0.22,1.59) |
| **Parents communicate about FP methods** |  |  |  |
| No | 87 (57.6) | 1 |  |
| Yes | 64 (42.4) | 1.47 (0.62-3.49) |  |
| **Money hindered use of FP** |  |  |  |
| Yes | 24 (16.3) | 1 |  |
| No | 123 (83.7) | 1.40 (0.51-3.81) |  |
| **Have a friend who used FP** |  |  |  |
| Yes | 93 (61.2) | 1 | 1 |
| No | 59 (38.8) | **0.40 (0.17-0.93)** | **0.31 (0.12,0.82)\*\*** |
| **Ever heard about FP** |  |  |  |
| Yes | 149 (97.4) | 1 |  |
| No | 4 (2.6) | 0.35 (0.06-2.00) |  |
| **Know where to get FP service** |  |  |  |
| Yes | 140 (92.1) | 1 |  |
| No | 12 (7.9) | 1.11 (0.24-5.27) |  |
| **Source of information about FP** |  |  |  |
| Health institution | 31 (20.7) | 1 |  |
| Mass media | 17 (11.3) | 0.55 (0.14-2.17) |  |
| Classroom | 23 (15.3) | 1.85 (0.33-10.42) |  |
| More than one | 79 (52.7) | 0.80 (0.27-2.36) |  |
| **Level where FP was taught** |  |  |  |
| Primary | 20 (13.2) | 1 |  |
| Secondary and above | 132 (86.8) | 1.80 (0.65-4.98) |  |
| **Awareness of FP availability at nearby facility** |  |  |  |
| Yes | 122 (80.3) | 1 | 1 |
| No | 30 (19.7) | **0.38 (0.16-0.90)** | 0.45 (0.16,1.25) |

**\*Adjusted for age (in years), family support for FP (yes/no), peer support for FP (yes/no) and awareness of FP availability at nearby facility**
**\*\* Adjusted for age (in years), family support for FP (yes/no) and awareness of FP availability at nearby facility**

### Factors associated with utilization of family planning methods

Among participants aged 19–21 years the odds of family planning use was 0.20 times lower compared to those aged 15–18 years (COR 0.20; 95% CI: 0.02,1.79). Moreover, the odds of family planning use among participants aged 22-24 years were 0.41 times lower compared to those aged 15-18 years (COR 0.41; 95% CI: 0.04,4.52). On the other hand, females had 2.84 times higher odds of family planning use as compared to males (aOR 2.84; 95% CI: (1.05,7.67)

(Table 2). During adjusted analysis, sex remained significantly associated with family planning use. Participants whose families support family planning use had 0.58 times lower odds of family planning use when compared to those whom their families do not support family planning use. However, the difference was not significant (aOR 0.58; 95% CI: 0.22,1.59). Moreover, those whose peers don’t use family planning had 0.31 times lower odds of family planning use as compared to those whose peers use family planning and the difference was significant in the multivariable analysis (aOR 0.31; 95% CI: 0.12,0.82).

The odds of family planning use among those who reported that family planning services are not available at the nearby facility were 0.38 times lower than those whom nearby facilities provide family planning services (cOR 0.38; 95% CI: 0.16-0.90) at this level it was significant. However, when adjusted with other factors it was found not to be significant (aOR 0.45; 95% CI: 0.16-1.25).

## DISCUSSION

This study aimed at determining the prevalence and factors associated with utilization of family planning methods among youth in Nyamagana District, Mwanza region. About 83.2% of all participants who have ever had sex used any modern family planning method, with condom being the widely used method. Reporting use of a modern family planning method was more likely among females. Peer pressure was an important factor in influencing use of a modern family planning method as participants whose peers did not use any FP method were also less likely to use a modern FP method. Although awareness on the availability of modern FP services at a nearby facility influenced utilization of FP when assessed on its own, it did not significantly influence use of FP when considered together with other factors.

Eight out of ten sexually active youth reported to have ever used a modern family planning method. We can compare the prevalence of FP use among youth with that of the National Demographic Health Survey of 2022 (7). The prevalence of the present study is higher when compared to the national prevalence for FP among girls aged 15 – 19 years and 20 – 24 years of 15.2% and 29.8% respectively. Results of the present study are also higher when compared to what has been reported in Tanzania’s regions of Kilimanjaro 51% (15), 57% in Dodoma (16) and other African countries of Malawi 60% and Namibia 61% (17, 18). However, these findings are a little closer to those reported by studies in Botswana of 75%, Eswatini 70% and Zimbabwe 65% (17, 18). Studies that reported results closer to what has been reported in the current study included both males and females while their counterpart enrolled only females. Reports of male condom being the most preferred method of FP partly collates higher FP use in the group. Also, differences in time of survey and efforts engaged by the government plus other stakeholders in fighting teenage pregnancy can explain the difference. Currently the government has revised the education laws by criminalizing sexual activity and marriage of school going girls (19). Also the government through the sexual offence act, criminalizes all teenage pregnancies among youths aged below 18 years and especially so for school going girls (20,21). In addition to these reviewers to laws, the government in 2021 strengthened the operationalization of school guidelines by lifting ban from return back to school of girls who were pregnant (22). All these efforts contribute towards increases use of FP among in school youths.

This study reported condom as the most commonly known and frequently used family planning method, the possible reason for this could be because condom can be easily accessed ranging from retail shop to pharmacy. A few participants mentioned other methods such as injectable, pills, implants, natural methods (calendar and withdrawal) and IUCD. This tallies with studies done in the country (23), Uganda (24), Sub-Saharan Africa (25) and in low-and middle-income countries (26) where condom was the most preferred method of FP.

The results from this study reported that females are more likely to report higher utilization of family planning methods. This results are similar with other studies conducted in the country (15,16,27), other East African Countries of Uganda (28), Kenya (29) and Sub-Saharan Africa (25,30), where sexually active female youth were found to practice contraception. This can be explained with the fact that, females, apart from fear of getting STIs like men, they are also afraid of getting pregnant because they may have to drop, end, interrupt or postpone their education (15,16,28) Furthermore, young women do engage in sexual activity with older men (inter-generational sex), of which men are afraid of being taken to court as a result of teenage pregnancy. The current reinforcement of school laws and the sexual offence act (19–22) further institutes this fear among these older men, making them more likely to use FP method during sexual activity with young women.

Peer pressure has been found to be an important factor associated with family planning methods use in the sense that those whose peers use contraception, are more likely to act the same. This finding is in line with other studies in the country (15,16) and from nearby countries (28–30). This can be reasonable with the circumstances that youth are more influenced with peer’s opinions in different issues. Peers create opportunity to exchange information and give out their sexual experiences including where they can access FP services. Taking this fact in consideration with the findings from this study which pointed out that awareness of FP availability at nearby health facilities significantly associated with family planning method use when it was assessed on its own, indicates the strength of peer to peer influence in accessing FP services. Peers are more likely to share with other peers where to source FP methods including how to access. A peer friendly source of FP methods will likely be popular among youth if peers did accept it when compared to other identified sources. In that regard, knowing a place where FP methods are available per see is not compelling to youth over peer influence. This was better described in a systematic review of studies on factors influencing access to and utilization of youth-friendly sexual and reproductive health services in sub-Saharan Africa (31). Ideally, peer-friendly source of FP works best with peer influence to drive high uptake of services among youth.

In the present study, it is worth noting that family support did not significantly influence the utilization of family planning methods. Although parental support has been shown to be a critical factor in other context, where it enhance adolescents’ confidence in understanding and meeting their reproductive goals, the situation was not similar with what has been shown in our present study Youth in the setting where this study may lack confidence in facing their parents regarding the use of family planning secondary to many reasons that presumably include uncertainty in their parent’s attitude towards family planning, cultural restrictions on discussing sexual related topics with their parents and failing to reveal to their parents that they are already sexually active. However, these reasons need further exploration using a qualitative approach.

### Strength and Limitation of the study

One of the main strengths of the present study was being able to enroll youth from both secondary school and colleges, making it possible to assess FP use within the whole spectrum of youth; 15 to 24 years of age. It provides evidence on improving utilization of family planning among youths, a key priority at present of the Government of Tanzania in achieving FP20230 commitments and Sustainable Development Goals. The study has some limitation to be considered when interpreting the results. First, the results of this study may not be generalized to the community of youths because it focused on the selected secondary schools and University College. However, the result may be generalized among school and university students in areas with similar setting. Secondly, due to its cross-sectional nature, cause and effect cannot be established because exposure and outcome data were collected simultaneously. Although data collection for this research was done several years ago, the results are still crucial in contributing towards addressing the low prevalence of utilization of family planning methods. Such low utilization rates call for more data in the country to understand factors influencing use of family planning. Lastly, since this study measured reported use of family planning method rather than the actual practice, prevalence of family planning use may be over-reported or under-reported. However, this was overcome by making responses anonymous, clarifying that the information gathered will be treated with strict confidentiality, will not be shared with teachers and can be used for study only.

### Conclusion

Satisfying sexual life among youth can be attained through addressing their sexual and reproductive health needs which is central in reducing maternal mortality caused by complications related with unwanted pregnancies. The reported prevalence of utilization of family planning methods in this study signifies that, there is high sensitization of FP method use among learned youths. Despite the fact that the study reported high prevalence of family planning use and peer pressure being the important factor, still there is a need for the provision of comprehensive family planning education at a much earlier level among youth population. Targeted strategies and initiatives about utilization of family planning methods among youth are required through a peer to peer approach.

### Recommendation

Peer pressure has been found to be an important factor in influencing use of FP among youth. The government and other stakeholders need to recognize this fact and improvise peer related approaches in scaling up the use of FP services among youth in similar setting. Further qualitative studies should be conducted to explore understanding on the role and dynamics around peer influence in relation to expand more reach in contraceptive use among youth.

## Data Availability

All relevant data are within the manuscript and its Supporting Information files.

## Acknowledgments

It’s our pleasure to thank the Kilimanjaro Christian Medical University College Research Ethics and Review Committee (CRERC), Nyamagana District Executive Officer, TANDABUI institute of health and allied sciences for permission granted to conduct this research. We would also like to acknowledge directors, staffs and students of schools participated in this study.

## Author’s contributions

All authors contributed towards the very beginning of proposal writing, data collection, analysis, report writing. The final manuscript was read, edited and approved by all authors.

## Disclosure

The authors declare no conflict of interest during this work.

## Notes

### Competing Interest Statement

The authors have declared no competing interest.

### Funding Statement

The author(s) received no specific funding for this work.

### Author Declarations

Ethical clearance for conducting the study was obtained from Kilimanjaro Christian Medical University College Research Ethics and Review Committee (CRERC) with certificate number 2233. Permission to conduct the study was requested and granted from Nyamagana District Executive Officer, and director from TANDABUI institute of health and allied sciences with information sent to participating schools. Written informed consent was obtained from every study participant. Confidentiality was highly observed by ensuring that the participants use code number instead of their names. The participants were well informed that they have freedom to refuse from participation to the study or refrain from answering certain question that doesnșt want to, without any penalty.

